# Evaluation of anti-CENP reactivity in samples exhibiting the centromere HEp-2 pattern, which is associated with a better prognosis within the limited cutaneous systemic sclerosis spectrum

**DOI:** 10.1101/2024.08.08.24311414

**Authors:** Gerson D. Keppeke, Diana Landoni, Cristiane Kayser, Pedro Matos, Larissa Diogenes, Jessica Keppeke, Silvia Helena Rodrigues, Luis Eduardo C. Andrade

## Abstract

**Introduction/Objectives:** Anti-centromere antibodies are associated with limited cutaneous systemic sclerosis (lcSSc) and a more favorable prognosis. The centromere HEp-2 pattern (AC-3) suggests the presence of antibodies against CENP antigens, mainly CENP-B and A. This study analyzed the clinical and demographic associations of anti-centromere antibodies in a cohort of patients exclusively with the lcSSc form of SSc. The frequency of CENP-B and CENP-A reactivity in samples with the AC-3 pattern was also evaluated.

**Method:** Samples from 38 lcSSc patients with AC-3 were evaluated for reactivity to CENP-B and CENP-A using line-blot and ELISA. Clinical data from 68 lcSSc patients (20 AC-3 and 48 Non-AC-3) were analyzed.

**Results:** Of the AC-3 samples, 84.2% and 81.6% were reactive against CENP-B and CENP-A, respectively, by line-blot, and 92.1% were positive for CENP-B by ELISA. Concordance for CENP-B reactivity between ELISA and line-blot was 78.9%. Reactivity to both CENP-B and CENP-A was found in 68.4% of AC-3 samples, while one sample was positive only for CENP-A. Overall, 97.3% of AC-3 samples were reactive to CENP-B, and all were reactive to either CENP-B or CENP-A. Clinically, interstitial lung disease (ILD) was less frequent in AC-3 patients compared to Non-AC-3 (10.5% vs. 54.2%; p=0.001). Other organ involvement frequencies were similar.

**Conclusions:** Among lcSSc patients, anti-CENP reactivity is associated with a less severe prognosis, with ILD being less frequent in AC-3-positive patients. In addition, anti-CENP-B was the predominant autoantibody in samples yielding the AC-3 pattern, but anti-CENP-A reactivity was also prevalent and exclusive anti-CENP-A reactivity was also observed.

## 1. INTRODUCTION

Systemic sclerosis (SSc) is a chronic heterogeneous autoimmune rheumatic disease characterized by high mortality and morbidity. This condition involves immune dysregulation, vasculopathy in small arteries and capillaries, and excessive collagen production, resulting in fibrosis of the skin and internal organs [1-5]. According to the extent of skin involvement, SSc can be classified into: a) Limited cutaneous SSc (lcSSc) that involves the face and the skin distal to the elbows and knees; b) Diffuse cutaneous SSc (dcSSc) that involves the face, chest, trunk, and the skin both distal and proximal to the elbows and knees; and c) Absent skin involvement (SSc *sine scleroderma*) [6]. It is also possible to classify SSC according to the presence of autoantibodies. Some autoantibodies are more associated with lcSSc, such as anti-centromere, anti-Th/To, and anti-PM-Scl, while others are more associated with dcSSc and multi-organ involvement, such as anti-topoisomerase I, anti-RNA polymerase III, and anti-fibrillarin (a comprehensive review can be found elsewhere [5]). Each of these autoantibodies is related to specific disease manifestations, what makes them valuable tools for estimating prognosis in a given patient [5]. Furthermore, the 2013 American College of Rheumatology and the European League Against Rheumatism (ACR/EULAR) improved the classification criteria for SSc by introducing a scoring system that includes clinical and laboratory elements [7].

Anti-centromere antibody is one of the most frequent in SSc [8]. However, these can also be found at lower frequencies in other autoimmune diseases, including Sjögren’s syndrome (SjS), primary biliary cholangitis (PBC), isolated Raynaud’s phenomenon, and overlap syndromes [9]. Nevertheless, they are considered highly specific for SSc (>90%) and have been reported to precede the onset of clinical disease by months or years [10]. In fact, the guidelines of ACR/EULAR indicate that the overall diagnostic sensitivity and specificity of anti-centromere antibody detected by indirect immunofluorescence assay on HEp-2 cells (HEp-2 IFA) was 31% and 97.4%, respectively, comparing with patients with other systemic autoimmune rheumatic diseases (SARD) [6, 11-13].

When tested by HEp-2 IFA, anti-centromere antibodies reveal a characteristic discrete speckled nuclear pattern scattered throughout in interphase cells and aligned at the chromatin mass on mitotic cells, compatible with the topography of the centromeres (Figure 1A). This pattern is classified as the AC-3 pattern according to the International Consensus on Antinuclear Antibody (ANA) Patterns (ICAP; www.anapatterns.org) [14, 15]. Structurally, the centromere is the region where condensed chromatin assembles to the inner and outer kinetochore to attach to the microtubules, which are responsible for chromosome segregation during cell division. Although there are many CENP proteins (CENP-A, -B, - C, -D, -E, -F, -G, H) in the kinetochore [16], CENP-B and CENP-A are the main autoantigens, as they are most consistently correlated with the AC-3 positive pattern on HEp-2 IFA observed in autoimmune patients [12, 17]. CENP-C is also the target of autoantibodies, but usually in association with antibodies to CENP-B or CENP-A [18].

**Figure 1.**
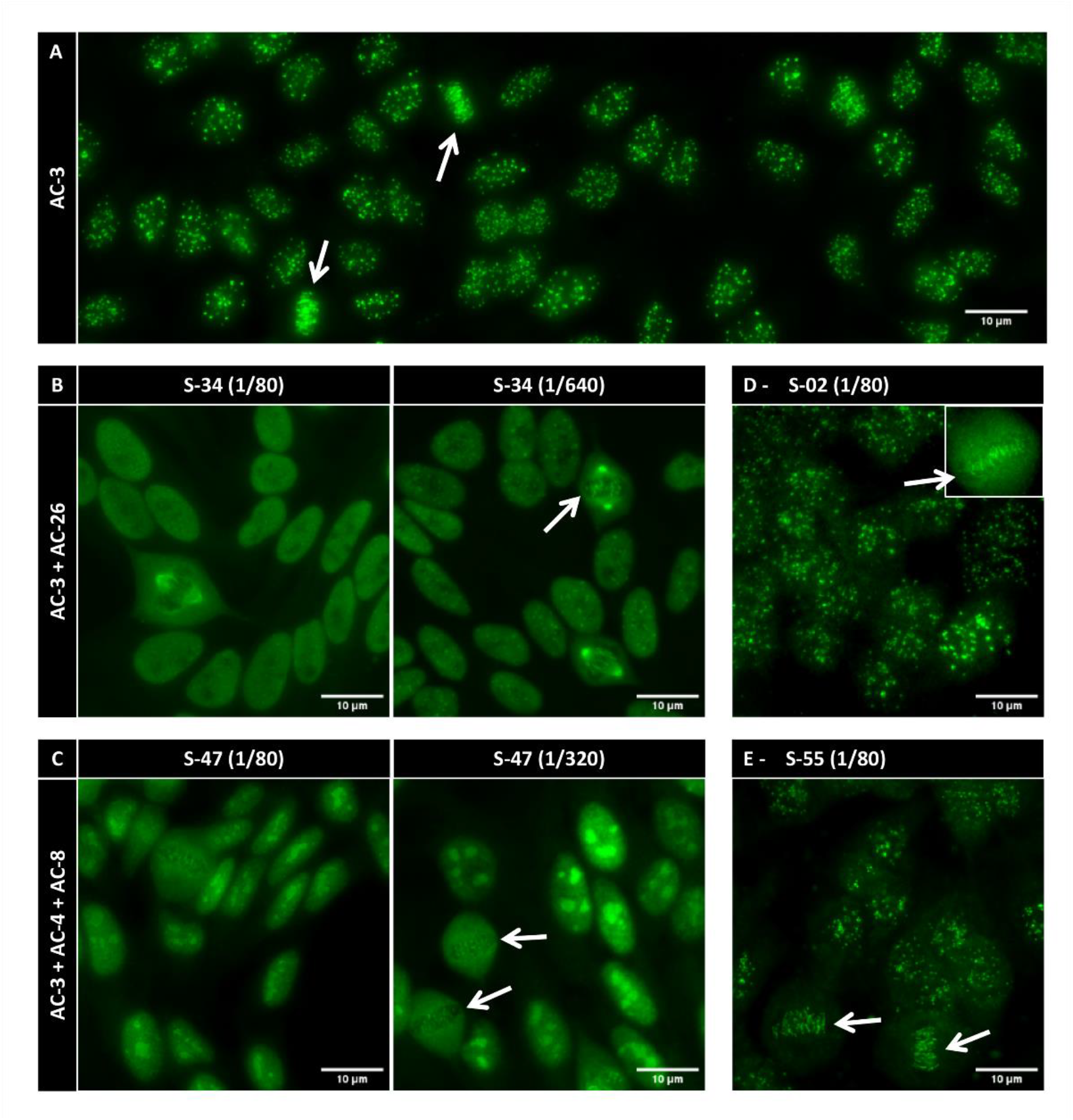
Representative images of the HEp-2 IFA for samples with the AC-3 centromere pattern. (A) The typical AC-3 pattern. (B) Sample S34 with multiple patterns combining the centromere AC-3 and the NuMA-like AC-26 patterns; the AC-3 became evidence at higher dilution. (C) Sample S47 with multiple patterns combining the nuclear fine speckled AC-4, the nucleolar homogeneous AC-8, and the centromere AC-3 patterns; the AC-3 became evidence at higher dilution. (D-E) Sample S02 had reactivity to CENP-B in the line-blot but not in the ELISA. Sample S55 was negative for CENP-B in both methods, but was positive for anti-CENP-A in the line-blot (Figure 3D). Arrows in all panels indicate the characteristic metaphase plate of the AC-3 pattern. Scale bar = 10 μm.

The 17kDa CENP-A and the 80kDa CENP-B share a cryptic linear epitope motif named G/A-PR/S-R-R mapped towards the C-terminal portion of CENP-B and the N-terminal charged region of CENP-A, which is the main epitope target of anti-centromere autoantibodies, [16, 19-21]. This may explain the nearly identical prevalence of reactivity to CENP-A and CENP-B in antigen-specific solid-phase assays among samples with the AC-3 centromere pattern in the HEp-2 IFA, leading some authors to suggest that ELISA could replace HEp-2 IFA, considering the level of expertise required for the HEp-2 IFA pattern analysis [12]. However, it is important to remember that HEp-2-IFA is a screening assay and does not provide the exact specificity for the nuclear antigen. Although the correlation of the AC-3 pattern with CENP-B/A autoantibodies is high, it is not flawless, especially if the sample produces multiple HEp-2 IFA patterns that may override the AC-3 pattern [5].

The HEp-2 IFA test, previously known as antinuclear antibodies (ANA), is a highly sensitive method for the screening for of anti-cellular antibodies (AC) [22]. The HEp-2 IFA provides information on the antibody serum concentration (titer) and possible autoantigen target (pattern). Various techniques, including ELISA, CLIA (chemiluminescent immunoassay), immunodiffusion, and immunoblotting, can be applied to detect specific antigen reactivity [5, 23]. Multiplex ELISA and dot/line-blots allow for the simultaneous testing of several autoantibodies. However these immunoassays usually use recombinant CENP-B or CENP-A proteins [12, 24], which could affect sensitivity, as demonstrated for other autoantibody systems [25]. Second-generation assays, like CytoBeads, combine IFA on HEp-2 cells and antigen-coated beads, creating a “2-in-1” solution for a one-step, two-level ANA test [26]. This approach may be useful for diagnosing patients who might not be detected with a negative HEp-2 IFA test but are positive for CENP-B by other methods. In general, CENP-B/A-specific immunoassays tend to show good agreement rates [27].

Most studies addressing the clinical associations of anti-centromere antibodies comprise general cohorts of SSc patients. Because anti-centromere antibodies are strongly associated with the lcSSc form of the disease, the clinical traits traditionally associated with anti-centromere antibodies are those that characterize lcSSc. Therefore, it is not well established how the anti-centromere antibodies correlate with the clinical spectrum of lcSSc. In this study, the clinical associations of anti-centromere antibodies were analyzed in a pure cohort of lcSSc patients. In addition, the anti-centromere reactivity in HEp-2 IFA (AC-3 pattern) was confronted with results of specific immunoassays for anti-CENP-B and anti-CENP-A antibodies.

### 1.1. Objective

We analyzed the possible clinical and demographic associations of anti-centromere antibodies in a cohort of patients exclusively with the lcSSc form of SSc. In addition, we evaluated the frequency of reactivity to CENP-B and CENP-A in samples with the AC-3 pattern on the HEp-2 IFA test.

## 2. METHODS

### 2.1. Patient samples

The patients were consecutively recruited from the Systemic Sclerosis Outpatient Clinic at Escola Paulista de Medicina, Federal University of Sao Paulo (UNIFESP), Brazil. Patients should meet the American College of Rheumatology/European League Against Rheumatism (ACR/EULAR) 2013 classification criteria for the limited cutaneous form of Systemic Sclerosis (lcSSc) [7]. In accordance with the Declaration of Helsinki, the patients signed an informed consent form to participate in the study and the research was approved by the Ethics Committee at UNIFESP (Plataforma Brasil CAAE: 59126320.1.0000.5505).

Demographic and clinical features were obtained from electronic medical records and reviewed by rheumatologists with expertise in SSc (C.K. and P.M.) as previously described [25, 28]. In brief, clinical data included age, sex, disease subtype, and disease duration (defined as the time between the first non-Raynaud symptom and the enrollment visit). Interstitial lung disease (SSc-ILD) was defined as the presence of interstitial abnormalities in chest high resolution computerized tomography (HRCT) and a forced vital capacity (FVC) on pulmonary function test lower than 80%. Pulmonary arterial hypertension (PAH) was considered in patients with group I PAH confirmed by right heart catheterization, according to previous established criteria [29]. Esophageal dysmotility was considered when confirmed in esophagogram or esophageal manometry.

The lcSSc patients were tested in the HEp-2 IFA test and subdivided into two groups according to the presence of the AC-3 pattern in the HEp-2 IFA test, respectively, the AC-3 group and the Non-AC-3 group.

### 2.2. Assays

The pattern and titer of the HEp-2-IFA were determined using commercial HEp-2 cell slides (#FA 1520-2010, Euroimmun; #51.100, AESKU), following the manufacturer’s protocol, with a 1/80 starting dilution and serial dilutions up to 1/2560. The slides were analyzed and images captured at 400x magnification using a fluorescent microscope (Axio Imager.M2, Carl Zeiss).

Anti-CENP-B reactivity was assessed using an indirect ELISA kit (#ORG 633, Orgentec), following the manufacturer’s protocol. A four-parameter logistic curve with four known concentration standards was applied (Figure 2B), and the interpolation of the samples’ optical density allowed the determination of anti-CENP-B reactivity in each sample in arbitrary units (U/mL). Samples with >10 U/mL were considered positive for anti-CENP-B, as recommended by the manufacturer. In addition, reactivity to CENP-A and CENP-B was determined by immunoblot (Euroline Systemic Sclerosis Nucleoli profile kit; Cat# DL 1532-6401 G, Euroimmun) following the manufacturer’s protocol (Figure 3). Although this kit can determine reactivity to other antigens, for this study we only considered reactivity to the CENP antigens. The manufacturer recommends interpretation of the line-blot result as: (−) negative; (+) one cross as *borderline*; (≥++) two crosses or more, as positive. Because one cross (*borderline*) may not represent true positives, as we have shown for other autoantigens [25], we considered positive samples only those with the immunostaining intensity ≥ two crosses (Figure 3).

**Figure 2.**
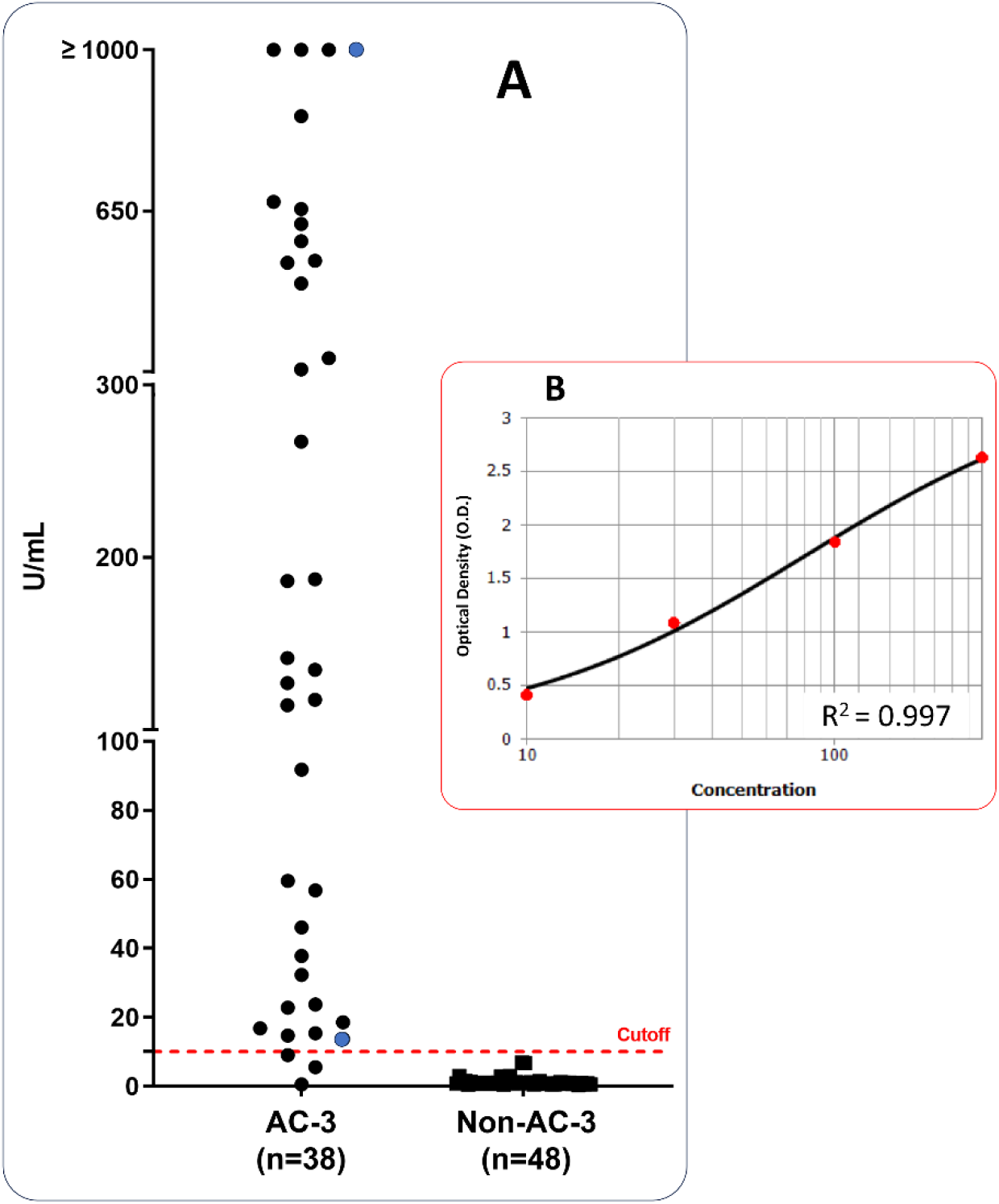
Reactivity to CENP-B in ELISA. (A) Anti-CENP-B reactivity tested by indirect ELISA. Distribution of anti-CENP-B reactivity in U/mL. The cutoff (red dotted line) was set at 10 U/mL as recommended by the manufacturer. The two data-points in blue indicate the two samples in which AC-3 was not initially reported, but it was observed in the HEp-2-IFA upon re-evaluation with serial dilution as detailed in Figure 1B and C, (B) A representative standard four-parameter logistic curve for the ELISA with anti-CENP-B standards.

**Figure 3.**
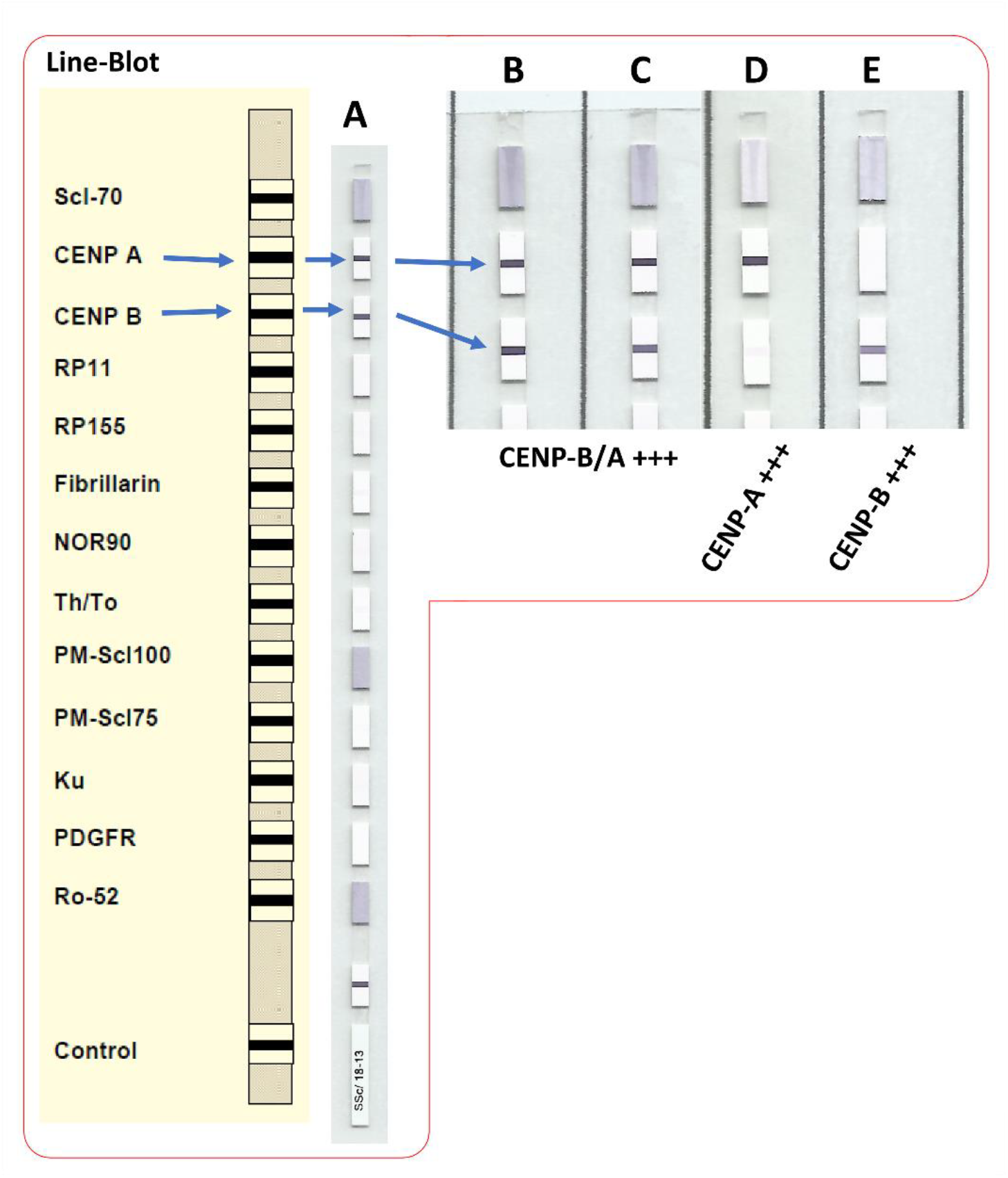
Anti-CENP reactivity in line-blot. (A-E) Euroline Systemic Sclerosis profile, (A-C) representative samples with 3 cross (+++) reactivity to anti-CENP-A and B, (D) representative sample with anti-CENP-A reactivity only, and (E) representative sample with anti-CENP-B reactivity only.

### 2.3. Data analysis

Immunofluorescence images were processed and panels assembled using ImageJ v1.53r software. Statistical analyses were performed using the software GraphPad Prism v7.0 or JASP v0.19.1. When comparing proportions, two-tailed Fisher’s exact test was applied. Quantitative and semi-quantitative parameters were accessed for normality distribution with Shapiro-Wilk test, followed by comparison with Mann-Whitney or Student t-test according to the distribution pattern. Correlations were evaluated with Spearman r test. P values were considered significant when below 0.05. Venn diagram was built with Venny 2.1 online tool.

## 3. RESULTS

There were 76 lcSSc patients, 38 classified into the AC-3 group and 48 classified into the Non-AC-3 group according to the presence of circulating anti-centromere antibodies. Concerning the AC-3 group, 29 (76.3%) patients showed a pure AC-3 pattern (Figure 1A) and nine (23.7%) patients showed a combination of the AC-3 pattern and other HEp-2 IFA patterns (Table 1). In this multiple pattern configuration, the centromere component tended to become more evident as the samples were further diluted (Figure 1B-C).

**Table 1.** Anti-CENP-B/A reactivity in 38 samples with the AC-3 pattern.

| HEp-2 IFA | Single pattern | Multiple patterns (AC-3 + others<sup>§</sup>) |  |
| --- | --- | --- | --- |
| AC-3 pattern | 29 (76.3%) | 9 (23.7%) |  |
| Titer range | 1/80 (n=1) to 1/2560 (n=6) |  |  |
| Median titer | 1/640 |  |  |
| Mean AC-3 titer (±SD) | 1/987 (±1/809) |  |  |
| CENP-B ELISA | Positives ≥10 U/mL | Negatives <10 U/mL |  |
| Proportions | 35 (92.1%) | 3 (7.9%) |  |
| Median reactivity | 141.5 | 5.5 |  |
| Mean reactivity (±SD) | 323.3 (±340.8) | 5.3 (±4.7) |  |
| Line-Blot | Positive (≥++) | Negative (-) and *Borderline* (+) |  |
| CENP-B | 32 (84.2%) | 6 (15.8%) |  |
| CENP-A | 31 (81.6%) | 7 (18.4%) |  |
| Correlation <sup>#</sup> | CENP-B ELISA | Line-Blot CENP-B | Line-Blot CENP-A |
| AC-3 pattern titer | 0.767*** | 0.432** | 0.419** |
| CENP-B ELISA | - | 0.594*** | 0.578*** |
| Line-Blot CENP-B | - | - | 0.676*** |

§ AC-4, AC-7, AC-8, AC-11, AC-21, AC-26. # Spearman r; \*\*p<0.01; \*\*\*p<0.001.

All samples were evaluated for anti-CENP-B reactivity in an indirect ELISA (Figure 2). Surprisingly, two samples that where originally not classified in the AC-3 group also tested positive, with reactivity above the cutoff of 10 U/mL (blue data-points in Figure 2A). These two samples (S34 and S47) were re-evaluated by serial dilution HEp-2 IFA and showed the discrete speckles at the metaphase plate typical of the centromere pattern at 1/640 and 1/320, respectively (arrows in Figure 1B and 1C). Consequently, we reclassified these two samples as containing more than one pattern, including the AC-3, and thus part of the AC-3 group (n=38). The AC-3 titer ranged from 1/80 to the highest dilution of 1/2560, with median of 1/640 and mean of 1/987 (Table 1).

As for the HEp-2 IFA pattern in the Non-AC-3 group, there were five negative samples (AC-0) and 43 with various patterns, such as nuclear fine speckled (AC-4; n=12), nuclear coarse speckled (AC-5; n=9), nucleolar (AC-8/9/10; n=15), DNA topoisomerase I (topo I)-like (AC-29; n=7), and miscellaneous patterns (AC-11, AC-18, AC-19, AC-21, AC-25; n=5), including six samples (14%) with more than one pattern.

Regarding the anti-CENP-B reactivity measured by ELISA, three (7.9%) of the 38 samples with AC-3 had results below the cutoff (Figure 2A) although all three samples had the AC-3 pattern at moderate intensity (titer 1/320; examples in Figures 1D and 1E). Therefore, 35 (92.1%) of the AC-3 samples were positive for anti-CENP-B by ELISA (Table 1).

Reactivity to CENP-B and CENP-A was also evaluated using line-blot assay (Figure 3). Most samples reacted with CENP-A and CENP-B (Figure 4) and one sample reacted only with CENP-A (Figure 3D). Seven samples reacted only with CENP-B (Figures 3E and 4). Altogether, the line-blot assay with the 38 AC-3 samples showed that 32 (84.2%) were reactive against CENP-B and 31 (81.6%) were reactive against CENP-A (Table 1 and Figure 4). From the three samples negative for anti-CENP-B antibodies in ELISA (Figure 2A), two were positive for anti-CENP-B in the line-blot assay and one (S55, Figure 1E) was positive only for anti-CENP-A (Figure 3D and Figure 4).

**Figure 4.**
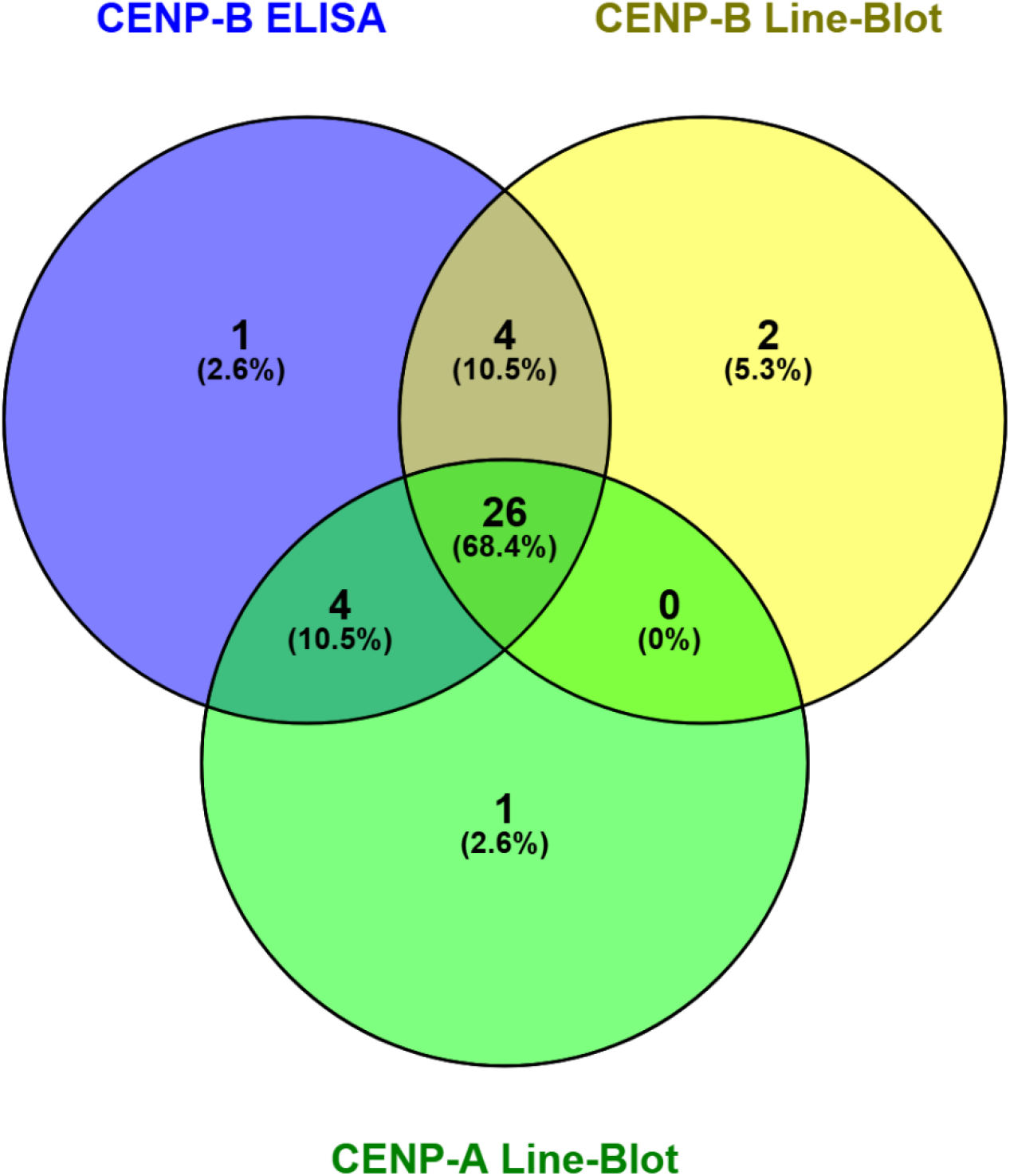
Venn diagram for anti-CENP-B/A reactivity in ELISA and line-blot assays in samples from the AC-3 group.

When comparing the reactivity to CENP-B in ELISA and line-blot assay, 30 (78.9%) samples were reactive against CENP-B in ELISA and line-blot methods (Figure 4). One sample was reactive against CENP-B only in ELISA and two samples were reactive against CENP-B only in line-blot.

Altogether, all 38 AC-3 samples (100%) showed reactivity against either CENP-B or CENP-A in at least one of the antibody-specific immunoassays. Only one sample was reactive exclusively against CENP-A, meaning that 37 (97.3%) were reactive against CENP-B in at least one method (Figure 4).

The correlation between the AC-3 titer in the HEp-2-IFA and the CENP-B reactivity in U/mL levels obtained in ELISA in the 38 AC-3 samples was high, r=0.767 (95% Confidence Interval 0.587-0.875; p<0.001). The correlation between the intensity of CENP-B reactivity in ELISA and the line-blot assay was satisfactory, r=0.594 (95% CI 0.330-0.772; p<0.001) (Table 1).

Clinical information was available for 20 patients from the AC-3 group (of whom 19 showed positive anti-CENP-B reactivity in ELISA and/or line-blot and one showed reactivity only to CENP-A) and for 48 patients from the Non-AC-3 group. The demographic data and clinical characteristics of these 68 lcSSc patients is depicted on Table 2. Patients in the AC-3 group were significantly older than those in the Non-AC-3 group but the duration of the disease was similar in the two groups. Regarding organ involvement, interstitial lung disease (ILD) was less frequently observed in patients in the AC-3 group (n=2, 10.5%) compared to those in the Non-AC-3 group (n=26, 54.2%; p=0.001), but the other parameters of organ involvement had similar frequency in the two groups (Table 2).

**Table 2.** Demographic and clinical features of the lcSSc patients according to the presence of anti-centromere pattern (AC-3) in HEp-2 IFA.

| Variable | AC-3 pattern<br>(n = 20) | Non-AC-3<br>(n = 48) | P |
| --- | --- | --- | --- |
| Age, mean ± SD (years) | 59.4 ± 12.5 | 50.5 ± 12.9 | 0.011 |
| Female, n (%) | 20 (100.0) | 41 (85.4) | 0.096 |
| Disease duration, mean ± SD (years) | 8.7 ± 6.3 | 6.6 ± 5.9 | 0.197 |
| *Organ involvement* |  |  |  |
| Digital ulcers, n (%) | 5 (26.3)* | 17 (35.4) | 0.571 |
| Esophageal dysmotility, n (%) | 14 (73.7)* | 38 (79.2) | 0.747 |
| Arthritis, n (%) | 6 (31.6)* | 19 (39.6) | 0.588 |
| FVC % of predict, mean ± SD | 84.5 ± 13.5 | 84.7 ± 19.9 | 0.967 |
| ILD, n (%) | 2 (10.5)* | 26 (54.2) | 0.001 |
| PAH, n (%) | 2 (10.5)* | 7 (14.6) | 1.000 |
| Cardiac involvement, n (%) | 0 (0.0) | 2 (4.2) | 1.000 |
| Scleroderma renal crisis, n (%) | 0 (0.0) | 2 (4.2) | 1.000 |

FVC: forced vital capacity; ILD: interstitial lung disease; PAH: pulmonary arterial hypertension.
\* Data available for 19 patients.

## 4. DISCUSSION / CONCLUSION

In this study, we analyzed the clinical and demographic characteristics of lcSSc patients according to the presence of anti-centromere autoantibodies and investigated the anti-CENP-B/A reactivity in samples displaying the AC-3 pattern in the HEp-2-IFA test. We showed that even among patients uniquely with the lcSSc subtype, the anti-centromere antibodies indicated a better diagnosis and less severe disease, as translated by a lower frequency of lung involvement. As expected, we confirmed the strong association between the AC-3 pattern and anti-CENP-B/A, as 100% of the AC-3 samples were reactive against CENP-B and/or CENP-A in at least one of the used immunoassays. Interestingly, however, the concordance rate between the solid phase assays themselves was weaker, as the agreement in anti-CENP-B reactivity between the ELISA and line-blot methods was only 78.9% as opposed to 100% concordance between HEp-2 AC-3 pattern and anti-CENP-B/A reactivity in solid-phase immunoassays. We also confirmed previous findings indicating that CENP-B is the dominant centromere autoantigen, as 37 (97.3%) of the AC-3 samples recognized CENP-B in at least one solid phase immunoassay whereas 31 (81.6%) AC-3 samples recognized CENP-A. In addition, among the 38 samples tested for antibodies to CENP-B and CENP-A, seven (18.4%) reacted exclusively with CENP-B and one (2.6%) was reactive solely against CENP-A. This result aligns with the concept that the AC-3 pattern in the HEp-2 IFA test is strongly associated with autoantibodies to CENP-B and/or CENP-A being CENP-B the dominant autoantigen [12, 18, 30].

The first publications describing the targets of autoantibodies that recognize centromeric antigens, namely the CENP proteins, as well as their association with lcSSc, date back almost half a century, resulting from studies conducted in Dr. Eng Tan’s laboratory in the early 1980s. At that time, lcSSc was classified by the presence of calcinosis, Raynaud’s phenomenon, esophageal dysmotility, sclerodactyly, and telangiectasia, collectively known as the CREST syndrome [31-33]. In SSc, as mentioned, the main autoantigens in samples with the AC-3 pattern are CENP-A and CENP-B [19]. Interestingly, the primary epitope on CENP-A, the G/A-PR/S-R-R motif, is also present on CENP-B and CENP-C. In fact, anti-CENP-A/B/C are frequently found in association in the same patient. It is important to note that the G/A-PR/S-R-R motif is not the only target, as these autoantibodies likely recognize other non-shared antigenic regions, providing strong evidence of intra- and intermolecular epitope spreading [20]. This is supported by our findings, where one patient showed reactivity only to CENP-A and seven showed reactivity only to CENP-B (and not to CENP-A), although we cannot rule out the presence of autoantibodies against other CENP antigens in these samples. Autoantibodies against CENP-A/B/C, as well as the less common CENP-D and CENP-E, and the very rare CENP-O [34], are all associated with lcSSc or the CREST syndrome [19, 20, 35]. CENP-D is primarily of the IgM type and tends to disappear over time [36]. Anti-CENP-E has been found in approximately 40% of patients with anti-CENP [37]. Autoantibodies against CENP-H are associated with Sjögren’s syndrome, particularly in patients without anti-Ro/La antibodies [38]. Finally, perhaps the most distinctive among the non-CENP-A/B antigens is CENP-F, a 330 kDa protein essential for cell cycle progression [39]. Anti-CENP-F antibodies are associated with various types of malignancies rather than SSc, PBC or Sjögren’s syndrome [40, 41]. These autoantibodies produce a different HEp-2 IFA pattern from the AC-3, referred to as the CENP-F-like pattern (AC-14) [39]. The presence of anti-CENP-F antibodies may serve as a marker for cancer [42].

Choosing the most appropriate method to determine anti-centromere antibodies is essential to ensure reliable results. In this study, no sample exhibited reactivity against CENP-B or CENP-A in the absence of the AC-3 pattern on the HEp-2 IFA. Conversely, all samples with the AC-3 pattern demonstrated reactivity against CENP-B/A in at least one assay. However, in some cases, the AC-3 pattern was visible only at higher dilutions and required a keen eye to identify the characteristic metaphase plate. Furthermore, we observed that some samples with the AC-3 pattern were negative in at least one of the solid-phase immunoassays. The concordance for CENP-B reactivity between ELISA and line-blot was less than 80%. Since clinical laboratories often use only one type of kit, they may fail to report samples with anti-centromere antibodies when relying solely on a solid-phase immunoassay. Thus, the data presented here provide additional evidence supporting the ACR/EULAR recommendation to use the indirect immunofluorescence assay on HEp-2 cells as the screening method for autoantibodies in rheumatic diseases, as commented elsewhere [43, 44], and to consider the reported pattern when interpreting solid-phase immunoassay results. Altogether, these findings indicate that HEp-2 IFA may be the most appropriate method of detection of anti-centromere antibodies.

Previous studies have demonstrated that anti-centromere autoantibodies display a less severe SSc disease and better prognosis [11, 30]. In fact, anti-centromere autoantibodies correlate with less frequent elevations in serum creatine kinase, digital ulcers, joint contractures, interstitial lung disease (ILD), scleroderma renal crisis, arthritis, and myositis, among others [12, 30]. However, these studies have inferred these associations in cohorts of patients with both forms of SSc, raising the possibility that the obtained associations are secondary to the primary association of anti-centromere antibodies to lcSSc, the more benign form of the disease. In our cohort constituted exclusively by lcSSc patients, we could confirm a lower frequency of ILD among lcSSc patients with anti-centromere antibodies compared with those without these autoantibodies. This finding suggests that the presence of anti-centromere antibodies further discriminate a subgroup of lcSS patients with more favorable prognosis. In a cohort comprising exclusively lcSSc patients, anti-centromere antibodies were associated with better prognosis and less severe disease. As proposed by a recent publication, individual autoantibodies associate with specific SSc characteristics [30]. Since ILD is the leading cause of death in SSc patients [45], our results suggest a less severe disease indicated by the less frequent ILD in SSc patients with anti-centromere autoantibodies.

In conclusion, lcSSc patients with anti-CENP reactivity appear to have a less severe prognosis, as ILD was less frequent in lcSSc patients with positive AC-3 pattern as compared to those with no anti-centromere reactivity. All samples with the AC-3 centromere pattern in HEp-2 IFA displayed reactivity to CENP-B or CENP-A in at least one of the applied tests, meaning the HEp-2 IFA method was 100% sensitive in detecting antibodies to CENP-A and CENP-B. One sample showed reactivity only to CENP-A, and of the 38 samples with AC-3, ∼82% were positive for CENP-A. Regarding CENP-B reactivity, ∼84% were positive by line-blot and ∼92% by ELISA, but only 30 samples were positive for CENP-B in both the ELISA and line-blot methods, with a concordance of ?80%. This means that anti-CENP-B is the predominant autoantibody in samples yielding the AC-3 pattern, but exclusive anti-CENP-A reactivity can also occur less frequently, as observed in only one sample in our cohort.

## Data Availability

All data produced in the present study are available upon reasonable request to the authors by qualified researchers.

## 5. DECLARATIONS

## Declarations of interest

The authors declare that they have no competing interests.

## Funding

This work was supported by Sao Paulo Government agency FAPESP (Sao Paulo State Research Foundation) grant numbers #2017/20745-1, #2021/04588-9 and #2023/17946-6, granted to GDK, LD and CK, respectively. Additionally, LECA was supported by the Brazilian research agency National Council for Research (CNPq), grant #PQ-1D 310334/2019-5. GDK also received support during the development of this work from Agencia Nacional de Investigación y Desarrollo de Chile (ANID), project No. [86220018].

## Ethics approval and consent to participate

In compliance with the Helsinki Declaration, the patients signed an informed consent form to participate in the study before donating their samples. The research was approved by Local Ethics Committee at the Federal University of Sao Paulo (Plataforma Brasil CAAE: 59126320.1.0000.5505).

## Acknowledgments

Parts of the manuscript (introduction/discussion) was originally written in Portuguese or Spanish, the mother language of the authors, and translated to English with the help of online tools such as ChatGTP with GPT-4-turbo (by OpenAI). The final text underwent proof by an English language expert, therefore, after using the online tools, authors reviewed and edited the content and take full responsibility for the content of the publication.

## Notes

### Competing Interest Statement

The authors have declared no competing interest.

### Author Declarations

Ethics committee of Federal University of Sao Paulo - Brazil gave ethical approval for this work (Plataforma Brasil CAAE: 59126320.1.0000.5505).

### Summary of Updates

Data presented on Table 1 and 2 were updated and reanalyzed. Consequently, parts of results were updated and the discussion/conclusions rewritten (updated) to present our conclusions clearly. References were also updated with the inclusion of 2 new references.

